# Scalable Incident Detection via Natural Language Processing and Probabilistic Language Models

**DOI:** 10.1101/2023.11.30.23299249

**Authors:** Colin G. Walsh, Drew Wilimitis, Qingxia Chen, Aileen Wright, Jhansi Kolli, Katelyn Robinson, Michael A. Ripperger, Kevin B. Johnson, David Carrell, Rishi J. Desai, Andrew Mosholder, Sai Dharmarajan, Sruthi Adimadhyam, Daniel Fabbri, Danijela Stojanovic, Michael E. Matheny, Cosmin A. Bejan

## Abstract

Post marketing safety surveillance depends in part on the ability to detect concerning clinical events at scale. Spontaneous reporting might be an effective component of safety surveillance, but it requires awareness and understanding among healthcare professionals to achieve its potential. Reliance on readily available structured data such as diagnostic codes risk under-coding and imprecision. Clinical textual data might bridge these gaps, and natural language processing (NLP) has been shown to aid in scalable phenotyping across healthcare records in multiple clinical domains. In this study, we developed and validated a novel incident phenotyping approach using unstructured clinical textual data agnostic to Electronic Health Record (EHR) and note type. It’s based on a published, validated approach (PheRe) used to ascertain social determinants of health and suicidality across entire healthcare records. To demonstrate generalizability, we validated this approach on two separate phenotypes that share common challenges with respect to accurate ascertainment: 1) suicide attempt; 2) sleep-related behaviors. With samples of 89,428 records and 35,863 records for suicide attempt and sleep-related behaviors, respectively, we conducted silver standard (diagnostic coding) and gold standard (manual chart review) validation. We showed Area Under the Precision-Recall Curve of ∼ 0.77 (95% CI 0.75-0.78) for suicide attempt and AUPR ∼ 0.31 (95% CI 0.28-0.34) for sleep-related behaviors. We also evaluated performance by coded race and demonstrated differences in performance by race were dissimilar across phenotypes and require algorithmovigilance and debiasing prior to implementation.

## Background

Incident detection refers to identifying new occurrence of relevant events from existing data assets and systems. Clinical examples of incident events include myocardial infarction, overdose from substance use, or suicide attempt. Precise detection at enterprise- or system-scale from healthcare records remains a major challenge. Once a product is Food and Drugs Administration (FDA) approved, post marketing safety surveillance for this medication includes both active processes like Sentinel and passive processes like the FDA Adverse Event Reporting System (FAERS).^1–4^ Identifying adverse drug events (ADEs) or new onset diseases that might relate to those new medications remains paramount.^5,6^ Outside of FDA regulatory processes, population health requires ascertainment of clinical incidents to allocate resources and properly support frontline staff, e.g., at triage in emergency settings.^7^ Precision medicine and predictive modeling initiatives (e.g., Clinical Decision Support [CDS]) also depend strongly on comprehensive ascertainment of phenotypes across biobanks and healthcare data repositories to power studies appropriately and minimize type I error.^8,9^

Significant attention has been given in guiding processes around reporting in post market surveillance,^10^ but gaps remain. Spontaneous reporting might be effective but requires awareness and understanding among healthcare professionals.^11^ It also has known limitations including under-reporting, duplication, and vulnerability to media attention or other trends in reporting such as sampling variation and errors or bias in reporting.^12^ Novel systems that leverage computational automation to achieve scalability might improve incident detection in healthcare broadly and post market surveillance specifically.

Another challenge for incident detection relates to the ability to determine whether the event started in the past, or *prevalent*, or whether it is a new occurrence, or *incident*. Many prevalent chronic conditions might have acute incident exacerbations, e.g., chronic obstructive pulmonary disease exacerbations, or be punctuated by incident clinical events, e.g., suicide attempts. Even when structured diagnostic codes exist for such events, differentiating whether those codes describe a new incident remains difficult. Current incident detection systems depend on structured data though efforts to expand inputs to unstructured data are underway.^13^ Coding data are driven by billing processes and are prioritized by clinical relevance and reimbursement rates. Codes are often linked to conduct of procedures or diagnostic testing, are not deterministic for the coded condition, and still might not always captured.^14–16^ Restrictions or rules in patients’ insurance that impact coding practices may also complicate incident assessment.

Temporality stands as another major obstacle to accurate incident detection.^17^ Many events have sequelae that result in similar or identical new data inputs.^18–20^ Sequencing healthcare events or, more importantly, evaluating potential causal links depends on establishing temporal order. Further, healthcare data might be recorded for a given patient at a later date (e.g., clinical documentation after billing) or outside of a “healthcare encounter”, as defined by most major vendor Electronic Health Records (EHRs).

A final challenge – most open healthcare systems do not have broad interoperability or data sharing to enable incident detection. Efforts to ameliorate this concern include national or payor systems, e.g., Veterans Health Administration or Kaiser Permanente, state-supported interoperability, e.g., New York’s Healthix,^21,22^ and vendor-led tools for common users of EHRs, e.g., EPIC Systems CareEverywhere. But while a patient suffering, e.g., a myocardial infarction (MI), at one health system might not have billing codes recorded in EHRs at another, that patient or their family would likely report the event to providers of care in another health system to ensure optimal clinical decision-making and healthcare communication. However, while providers are expected to obtain and summarize relevant patient care leading up to an encounter or interaction, structured codes from prior care are not generally imported. A summary of outside care might be reliably documented in unstructured clinical text in the routine practice of medicine.

Natural language processing (NLP) permits extraction and detection of incidents from unstructured textual data, and has been used in event detection and disease onset before.^16,19^ It has also been applied to accurately identify social determinants of health to better understand the prevalence of these problems both within^23^ and across^24^ health systems including in FDA-linked initiatives like Sentinel.^25,26^ In work motivating this study, NLP has been applied to suicidal ideation and suicide attempt yielding accurate and precise ascertainment from unstructured text data agnostic to source data or EHR.^27^

To improve scalable incident detection using unstructured healthcare data, we developed and validated a novel incident phenotyping approach using unstructured clinical textual data agnostic to EHR and note type. It’s based on a published, validated approach used to ascertain social determinants of health and suicidality across entire healthcare records (prevalence). To demonstrate generalizability, we validated this approach on two separate phenotypes that share common challenges with respect to accurate ascertainment: 1) suicide attempt; 2) sleep-related behaviors. They have the additional rationale that identifying them might warrant further investigation and/or have regulatory relevance.

## Methods

### Cohort generation

Data were extracted from the Vanderbilt Research Derivative, an EHR repository including Protected Health Information (PHI), for those receiving care at Vanderbilt University Medical Center (VUMC).^28^ PHI were necessary to link to ongoing operational efforts to predict and prevent suicide pursuant to the suicidality phenotypic work here.^29,30^ Patient records were considered for years ranging from 1998 to 2022. For both suicide attempt and sleep-related behaviors, we focused on adult patients aged over 18 years at the time of healthcare encounters with any clinical narrative data in the EHR.

While the technical details of the Phenotypic Retrieval (PheRe) system adapted here have been published elsewhere,^23,27^ the algorithm’s retrieval method determined which records were included in this study. In brief, after query formulation to establish key terms for each phenotype (see “Automatic extraction…” below), this algorithm assigned scores to every adult patient record. To be included in this study, those records with any non-zero NLP score, i.e., any single term in the query lists, were included in subsequent analyses.

### Phenotype Definitions

Our team has published extensively in the suicide informatics literature on developing, validating, and deploying scalable predictive models of suicide risk into practice. As a result, suicide attempt was defined based on prior work using published diagnostic code sets for the silver standard^27^ and domain knowledge-driven definitions for the gold standard annotation (see below for details on both).

For sleep-related behaviors, our team reviewed the literature on methods to ascertain such behaviors from structured diagnostic codes. We also consulted with clinical experts in sleep-related behaviors and sleep disorders in the Department of Otolaryngology at VUMC (see Acknowledgement). This expertise informed both the silver and gold standards for this phenotype. These standards will be detailed below; in brief, the silver standard was hypothesized to be less specific and less rigorous a performance test than the gold standard yet easier to implement since it relied on structured data.

### Temporality

A major difference between our prior published NLP to ascertain phenotypes from EHRs relates to prevalence versus incidence.^23^ In prior work, we applied NLP to ascertain evidence of any suicide attempt or record of suicidal ideation from EHRs across entire healthcare records.^27^ In this work, the intent was to ascertain evidence of new, clinically distinct incidents of these phenotypes. The team discussed numerous potential temporal windows to focus the NLP including: i) healthcare visit episodes; ii) set time-windows, e.g., twenty-four-hour periods; iii) combinations of those two, e.g., clinical episodes plus/minus a time-window to capture documentation lag.

After discussion and preliminary analyses, we selected a twenty-four-hour period, midnight to the next midnight, as the window for this incident detection NLP approach. This window was chosen for clinical utility, simplicity, and agnosticism to vendor EHR or documentation schema. Operationally, this meant that we considered all the notes of a patient on a given day to encode a potential incident phenotype.

### Automatic extraction of phenotypic profiles from clinical notes

We developed a data-driven method to extract relevant text expressions for each phenotype of interest (see Figure 1). The method involved processing large collections of clinical notes from EHRs (including tokenization and extraction of n-gram representations such as unigrams and bigrams) and unsupervised training of Google’s word2vec^31^ and transformer-based NLP models such as Bidirectional Encoder Representations from Transformers (BERT)^32^ to learn context-independent and context-sensitive word embeddings. The extraction of phenotypic profiles consisted of iteratively expanding an initial set of high-relevant expressions (also called ‘seeds’) such as *‘suicide*’ as follows. First, we ranked the learned embeddings by their similarity to the seed embeddings. Then, we manually reviewed the top ranked expressions and selected the relevant ones as new seed expressions. The final sets of text expressions corresponding to each phenotype of interest are listed in eSupplement.

**Figure 1:**
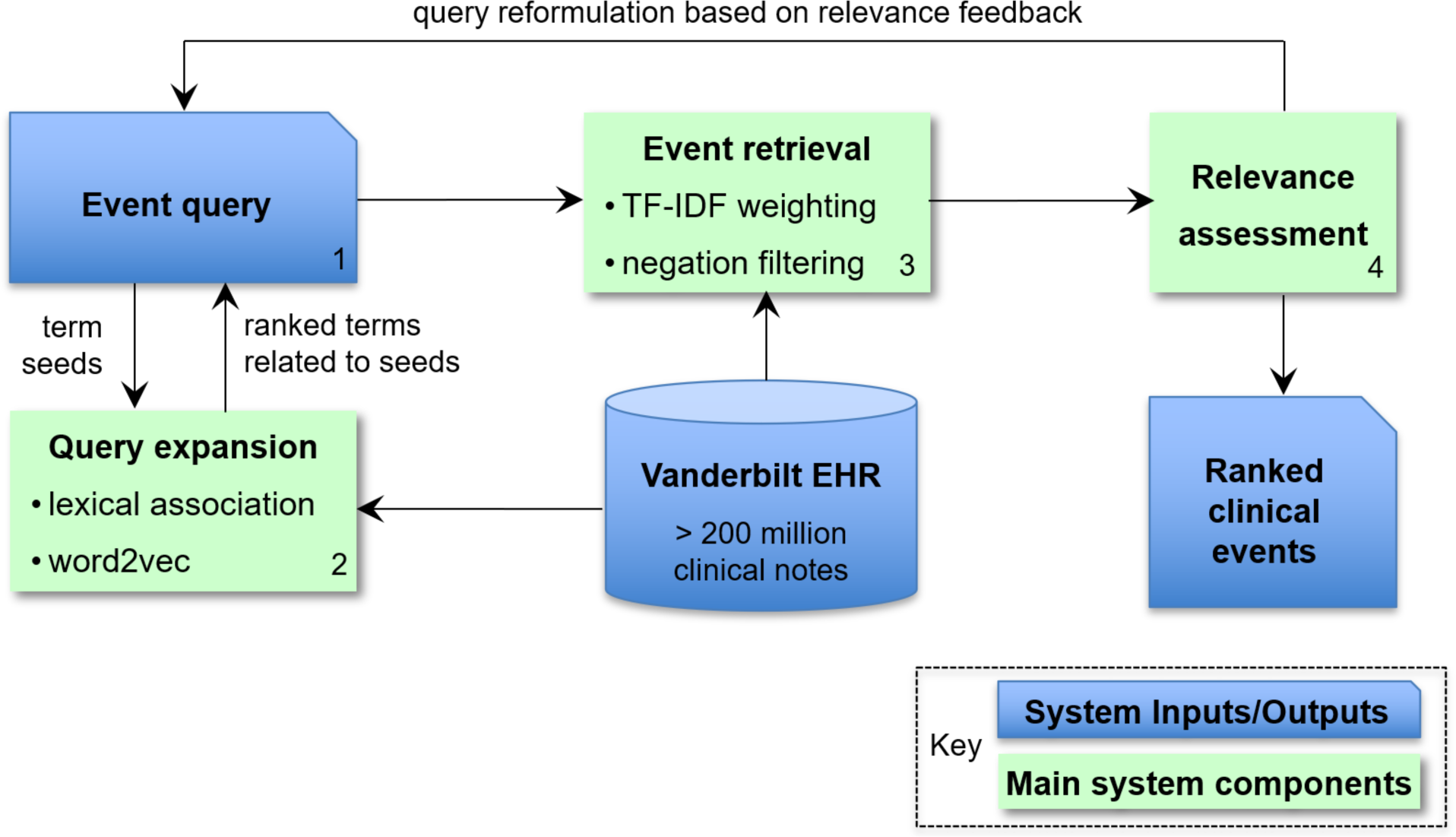
Overview of Automatic Extraction Process Enabling Incident Detection, Steps Numbered and Legend Shown.

### Large-scale retrieval of incident phenotypes

We implemented a search engine to identify incident phenotypes in all the notes from the Vanderbilt Research Derivative and to rank them by relevance to their profile. In this context, each phenotypic profile corresponds to an input query for the search engine while each meta-document comprising of all the notes of a patient on a given day encodes a potential incident phenotype. In the implementation framework, we represented the meta-documents and input queries as multidimensional vectors, where each vector element is associated with a single- or multi-word expression from their corresponding phenotypic profile. The relevance of a patient meta-document to a phenotype was measured as the similarity between their meta-document and input query vectors using the standard term frequency-inverse document frequency (TF-IDF) weighted cosine metric. The final NLP-based score was a continuous value ranging from low single digits (< 10) to hundreds (< 500 typically). Higher scores indicated more similarity and therefore more evidence of the phenotype.

To further improve the performance of our search engine, we performed query reformulation based on relevance feedback by iterative assessment of the top 20 retrieved incident phenotypes of each run.^23^ The selection and ranking of the incident phenotypes was performed using the Phenotype Retrieval (PheRe) software package in Java, which is available at https://github.com/bejanlab/PheRe.git.

### Silver standard generation

A silver standard represents a source of truth that might be less precise or more error-prone than ground truth, a gold standard. An advantage to silver standards remains their relative efficiency to generate and validate compared to more labor-intensive gold standards. To generate silver standards for sample size calculations for all phenotypes, we used ICD9CM (Clinical Modification) and ICD10CM diagnostic code sets to generate preliminary performance for the NLP to identify presence/absence of structured phenotypic data (i.e., International Classification of Disease [ICD] codes).^33^ For suicide attempt, we used validated code sets from published literature.^29,34^ For sleep-related behaviors, we reviewed the literature and adapted code sets from the literature with clinical experts in Sleep Medicine at VUMC. Codesets for all phenotypes used in this project are available in the eSupplement.

To evaluate preliminary performance, we used presence/absence of a single ICD code from the validated lists within thirty days of the date of NLP score calculation as a positive label (label = 1) and the absence as a negative label (label = 0). Thirty days was chosen as a common time period in which clinical risk is close enough to warrant documentation and intervention but not so close as to be imminent and required emergent care. The continuous NLP scores were a univariate ‘predictor’ of presence/absence. Performance was measured with typical information retrieval and model discrimination metrics: Area Under the Receiver Operating Characteristic (AUROC), Precision (P), Recall (R), P-R Curves, and F-measures (F1-score).

### Gold standard generation

The intent in gold standard generation was to generate corpora of charts across all NLP score bins (e.g., NLP scores from 5-10, 10-15, 15-20, …) to evaluate performance of the NLP incident detection system. Unlike top-K precision used in information retrieval, we sought insight into performance with confidence intervals across score spectra to plan for downstream CDS applications.

For sample size calculation, we used preliminary performance from the silver standards to calculate numbers of chart-days to extract for multi reviewer chart validation. The rationale for this key step was the need for 1) efficient chart review within a selected marginal error and 2) intent to understand NLP performance for all possible scores - not simply performance in the highest ranked charts, aka “top-K” performance typical in this literature where K is some feasible number of highly ranked records, e.g., K=200. To determine the number of encounters for chart review, we used the marginal error method which involves the half-width of the 95% confidence interval for the performance metrics in the investigated cohort. The process involved the following steps: (1) dividing the predicted risk scores into 5-point intervals and calculating the number of encounters in each interval; (2) estimating the precision and recall for each interval using ICD9/ICD10 codes; and (3) computing the sample size by setting the marginal error for the probability estimate in the individual interval to 0.05. We assumed that the number of positive encounters selected for chart review approximates a normal distribution and follows a hypergeometric distribution. The larger sample size between the calculated sample sizes for precision and recall estimates determines the required sample size for chart review.

To conduct chart review, annotation guides were developed and revised after initial chart validation training (fifty chart-days for each phenotype). These guides included instruction on labeling and factors contributing to label decisions. All reviewers (K.R., J.K., C.W. [adjudicator]) were trained on the required annotation and participated in a Training phase using fifty chart-day examples for each phenotype. Chart labels included: Positive – phenotype documented in notes “reports suicidal ideation”; Negative - phenotype documented as not present, e.g., “denied suicidal ideation”; Unknown – insufficient evidence to determine labels. In subsequent analysis after the third reviewer adjudicated disagreement and unknown labels, these three labels were collapsed into two: Positive; Negative. Annotation guides are available as eSupplements.

### Evaluation Metrics

Metrics to evaluate NLP performance mirrored those used in preliminary analyses above including P-R Metrics and curves; F1-score. We also calculated error by score bin to understand how well the NLP score performed across all thresholds. The intent was to replicate a common clinical implementation challenge – discretizing a continuous output from an algorithm into a binary event, e.g., a decision or an intervention that cannot be discretized in practice.

## Results

### Baseline patient characteristics by phenotype

Across both suicide attempt and sleep-related behaviors, the study cohorts included 89,428 and 35,863 patients, respectively. As outlined in Cohort generation above, these numbers included any records with at least one query term match in the day of notes for that patient. Baseline study characteristics at each patient’s first documented visit are shown (Table 1).

**Table 1:**
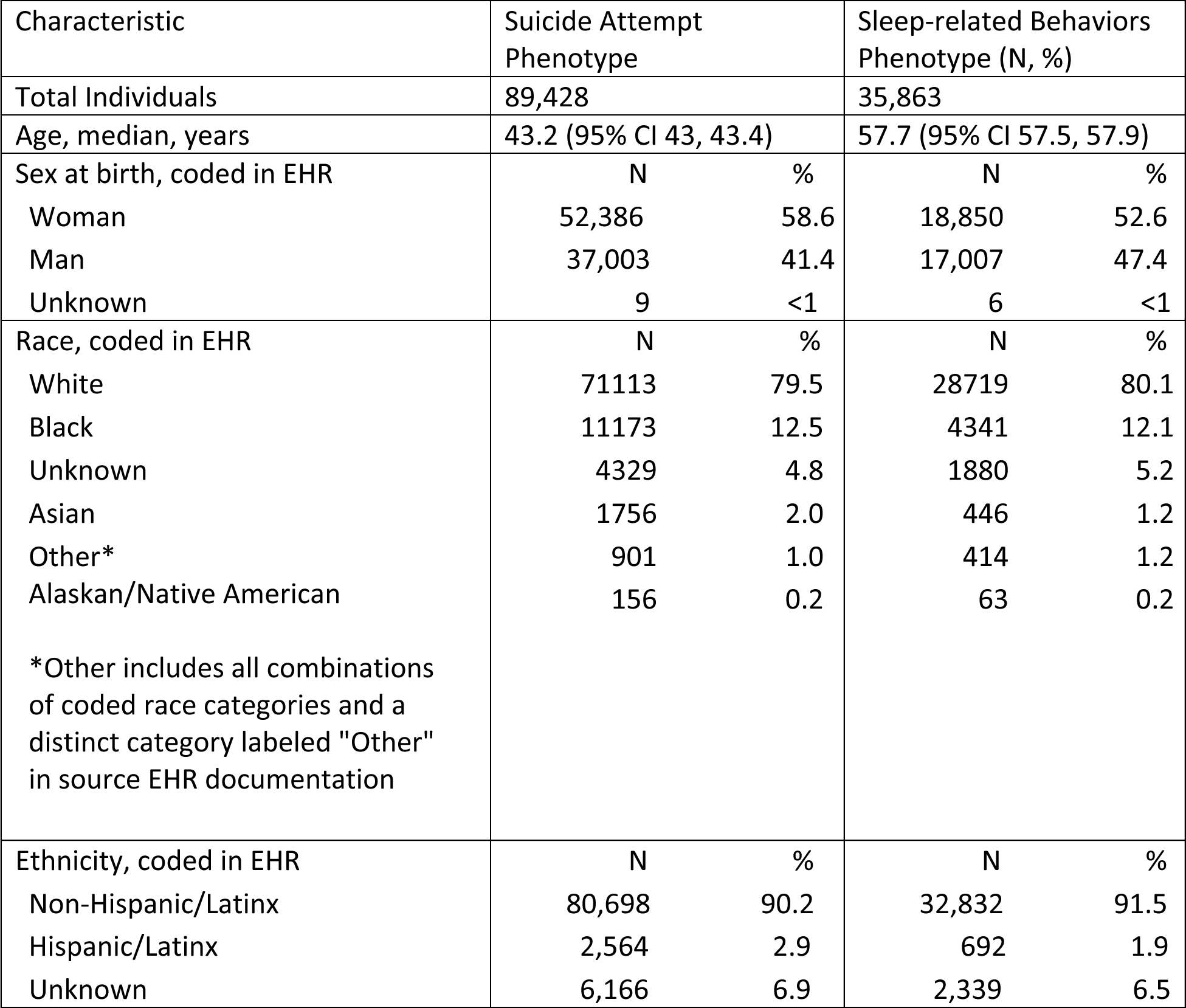
Baseline Study Characteristics.

| Characteristic | Suicide Attempt Phenotype |  | Sleep-related Behaviors Phenotype (N, %) |  |
| --- | --- | --- | --- | --- |
| Total Individuals | 89,428 |  | 35,863 |  |
| Age, median, years | 43.2 (95% CI 43, 43.4) |  | 57.7 (95% CI 57.5, 57.9) |  |
| Sex at birth, coded in EHR | N | % | N | % |
| Woman | 52,386 | 58.6 | 18,850 | 52.6 |
| Man | 37,003 | 41.4 | 17,007 | 47.4 |
| Unknown | 9 | <1 | 6 | <1 |
| Race, coded in EHR | N | % | N | % |
| White | 71113 | 79.5 | 28719 | 80.1 |
| Black | 11173 | 12.5 | 4341 | 12.1 |
| Unknown | 4329 | 4.8 | 1880 | 5.2 |
| Asian | 1756 | 2.0 | 446 | 1.2 |
| Other* | 901 | 1.0 | 414 | 1.2 |
| Alaskan/Native American | 156 | 0.2 | 63 | 0.2 |
| *Other includes all combinations of coded race categories and a distinct category labeled "Other" in source EHR documentation |  |  |  |  |
| Ethnicity, coded in EHR | N | % | N | % |
| Non-Hispanic/Latinx | 80,698 | 90.2 | 32,832 | 91.5 |
| Hispanic/Latinx | 2,564 | 2.9 | 692 | 1.9 |
| Unknown | 6,166 | 6.9 | 2,339 | 6.5 |

### Silver standard performance (diagnostic code classification)

From prior work and multiple validation studies of the silver standard diagnostic coding for suicide attempt, we used 58% PPV for ICD9 and 85% for ICD10 versions of these codes as the initial benchmarks.

For sleep-related behaviors, we had no similar benchmarks from prior work or the published literature. Using the presence of an ICD9 or ICD10 code for sleep-related behaviors, we note the NLP algorithm had a PPV of 60% to predict the silver standard and establish a benchmark for sample size calculation and chart review.

### Gold standard performance (chart validation)

Precision-recall performances of the NLP incident detection systems are shown including AUPR∼ 0.77 (95% CI 0.75-0.78) for suicide attempt and AUPR ∼ 0.31 (95% CI 0.28-0.34) for sleep-related behaviors.

P-R curves are shown with respect to coded race and smaller differences are noted for suicide attempt by coded race compared to sleep-related behaviors (Figure 2). Notably, the confidence intervals in the legend also overlap for both phenotypes (Figure 3).

**Figure 2:**
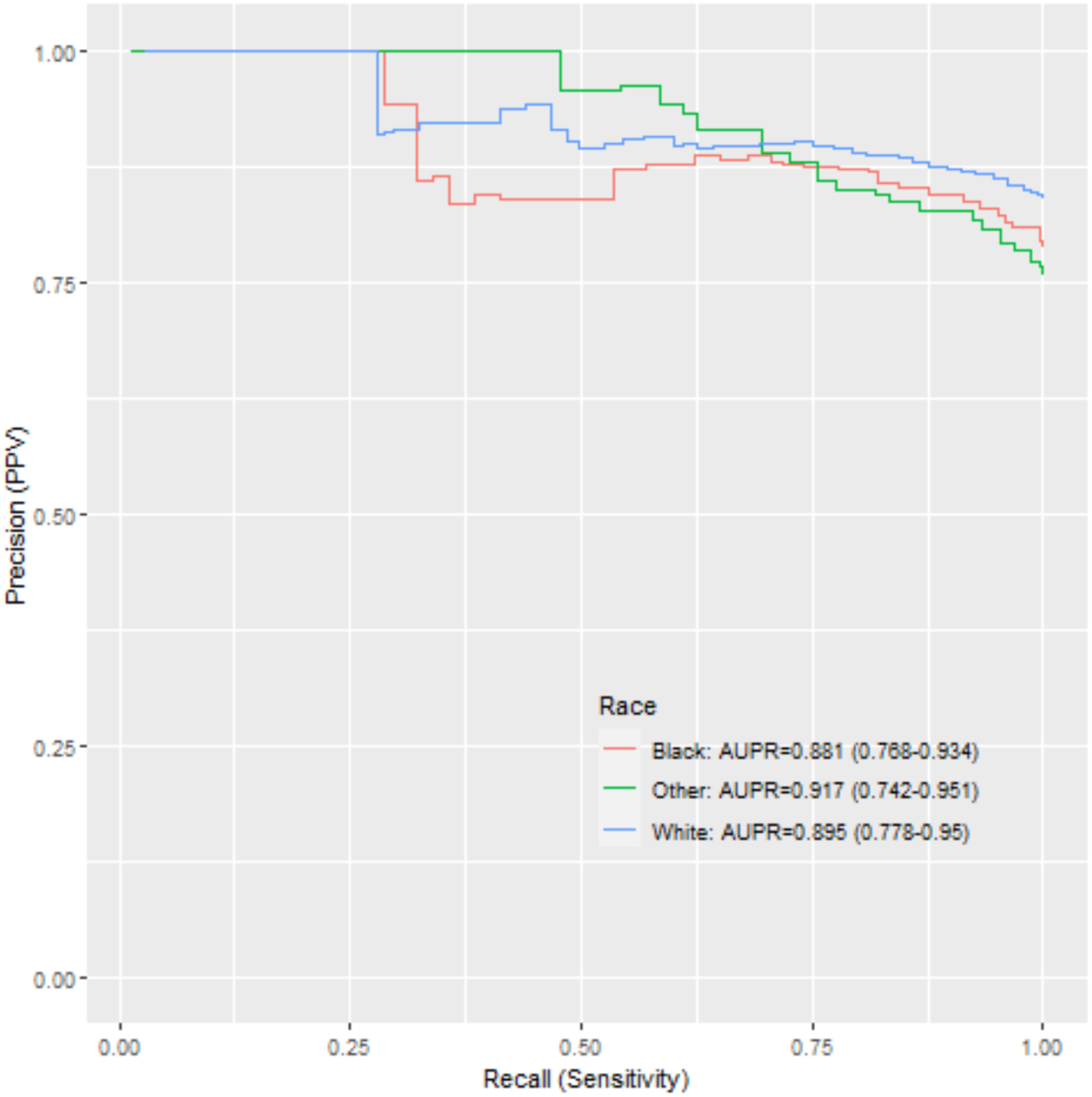
Precision-Recall of Suicide Attempt NLP Incident Detection by coded race.

**Figure 3:**
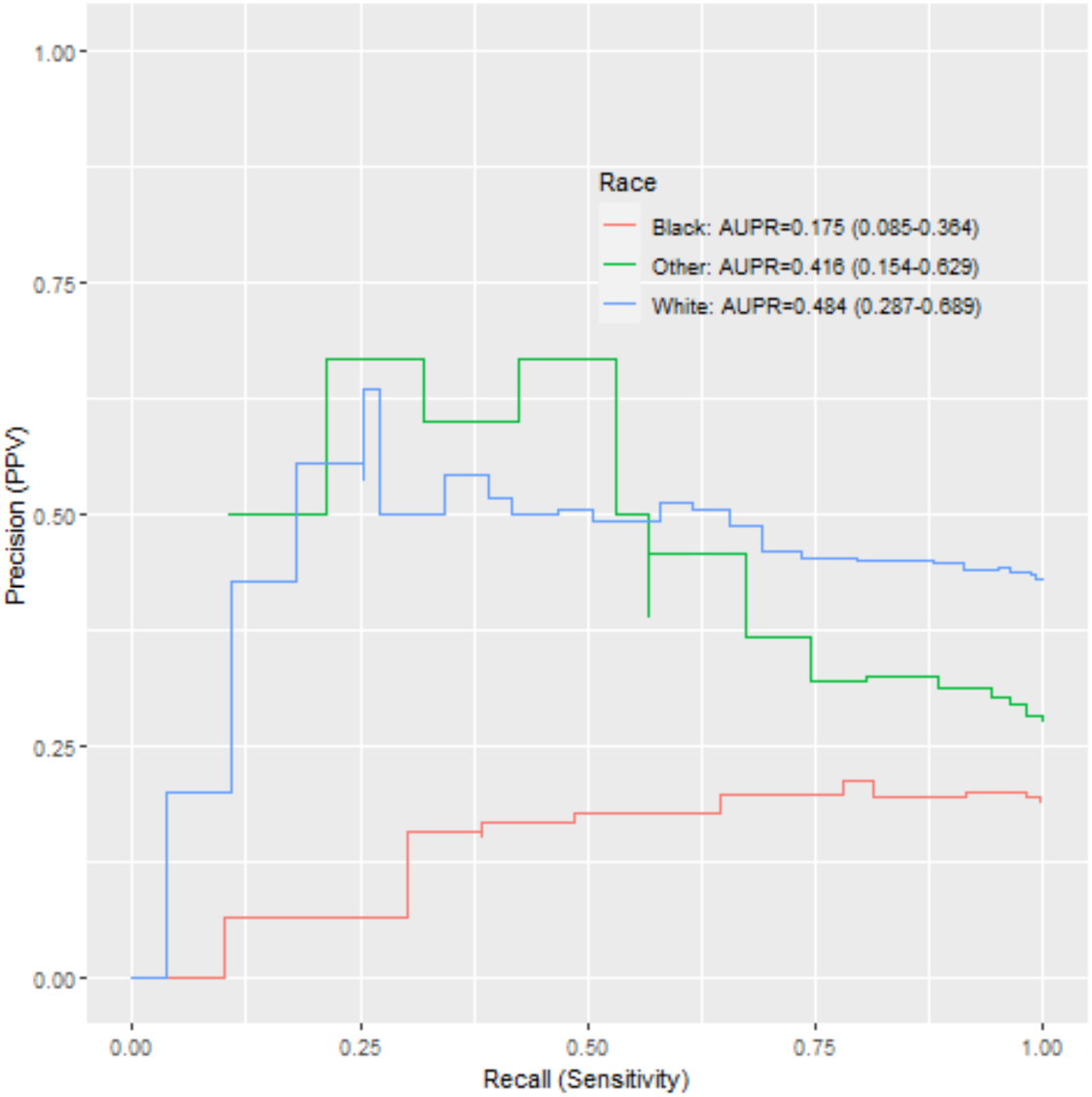
Precision-Recall of Sleep-related Behaviors NLP Incident Detection by coded race.

### Threshold Selection, Steps toward Implementation

Selecting a threshold for a hypothetical implementation where precision matters more than recall, we use the maximum F1-score. For suicide attempt NLP-based detection, for example, and our overall P-R performance above, an NLP score of 25 and higher would have an F1-score of 0.75 associated with a recall of 0.93 and a precision of 0.63.

Similarly calculated for sleep-related behaviors, the optimal F1-score-based threshold is 35 or above in which the F1-score would be 0.42, precision 0.33, recall 0.57. We note that this score, while the optimal model performance, might not represent an acceptable or optimal performance for specific applications e.g., defining an endpoint of interest in medical product safety surveillance.

## Discussion

In this study, an NLP-based incident detection system was developed and validated across two challenging and disparate phenotypes. Such detection was feasible to conduct using a twenty-four-hour period of documentation, agnostic to EHR architecture or data standard, or to underlying textual source systems. The implications of this system for initiatives like FDA Sentinel indicate scalable detection would be achievable with appropriate evaluation and milestones in the implementation path. For example, in these two phenotypes, gold standard manual chart review was necessary and would remain necessary for novel phenotypes on a subset of charts in development sites. But here as in many phenotypic examples, prior work facilitated the sample size calculation and chart review itself. Moreover, silver standard validation permitted efficient sample size calculation and error estimation across all possible NLP scores, not solely those highest ranked “top-K” records as in traditional information retrieval. Generally, teams experienced with these phenotypes would be well-suited to contribute to evaluating their accuracy before deployment. Even with imperfect coded race variables, performance differences were easily identifiable here. Algorithmovigilance^35^ to perpetuating or worsening disparities remains paramount in the evolution of systems like this one – not solely at initial algorithm validation but throughout the life cycle.^36^

Performance varied by phenotype with better performance with a phenotype that was more common and with a clearly defined clinically observable set of attributes in suicide attempt, though it remains challenging as a common label for a clinical event that takes many different forms at the individual level. Sleep-related behaviors, even when focused on sleepwalking, - eating, -driving, are still a group of diagnoses that may or may not be documented clearly in every visit even when present. That is, these selected phenotypes differ in clinical specificity and in the degree to which care will focus on them if observed. However, both might be associated with regulatory action if a new medication or device were shown to cause them. Incidence rates of a target phenotype remain key factors informing the development of predictive models like these.

Active efforts including a recent National Academy of Science, Engineering, and Medicine (NASEM) report suggest excluding race from genetic and genomic studies.^37^ Race remains a social construct with potential to reflect or worsen healthcare disparities if not handled appropriately. Use of clinical language does vary by race and this usage remains an important limitation of studies like this one. Here, coded race was used as a means of testing these NLP algorithms for disparate performance as an early checkpoint in model validation. Our findings are hypothesis-generating with respect to reasons for performance differences and more detailed analyses with better quality race data are indicated in any case. A parallel effort in replicating an approach like this one in new phenotypes would necessitate careful consideration of factors like demographics, clinical attributes or other that might undermine the successes of an incident detection system at scale.

Silver standard evaluation alone was not sufficient to estimate final NLP performance observed here. Both sleep-related behaviors and suicide attempt were associated with ∼ 60% PPV in silver standard, ICD-based, performance but suicide attempt as a phenotype was much better identified in this method than sleep-related behaviors. Thus, some manner of gold standard evaluation is indicated for adding new phenotypes to this system. To that end, annotation guides and annotator training facilitated rigorous multireviewer chart validation as did sample size calculation of numbers of required charts to review based on the silver standard.

This work builds on the work of others by adding to understanding of unstructured data-based phenotyping algorithms in neuropsychiatric phenotypes with emphasis on temporality and incident detection. Determining temporal onset of symptoms with NLP has been attempted in clinical areas including psychosis,^38^ perinatal health,^39^ and hematology.^40^ Deep learning has been used with NLP-based features to identify acute on chronic exacerbations of chronic diseases such as hypoglycemic events in diabetes.^41^ While phenotyping in suicidality has included NLP in numerous studies including those of this team, phenotyping in sleep disorders has been less commonly reported.^42,43^ This study adds to evidence that sleep-related behaviors might be less well-coded and well-documented than other neuropsychiatric phenotypes and therefore NLP-based algorithms to detect them were more challenging to develop.

## Data Availability

Because data include sensitive PHI, data are not available for dissemination outside the study team.

## Acknowledgements

Our team thanks Dr. David Kent in the Department of Otolaryngology at VUMC for expertise in sleep-related behaviors and insight into phenotypic definitions and acceptable silver standard diagnostic codes used here.

We thank Dr. Patricia Bright for reviewing our manuscript prior to submission.

## Funding

All investigators were supported on FDA WO2006. Dr. Walsh is also supported in part by NIMH R01MH121455 and R01MH116269.

Funders played no role in design and conduct of the study; collection, management, analysis, and interpretation of the data; preparation, review, or approval of the manuscript; and decision to submit the manuscript for publication.

## Author Contributions

Design and conduct of the study (all authors)

Data collection, management, analysis (CW, DW, QC, CB)

Chart validation (JK, KR, CW)

Interpretation of the data (all authors)

Preparation, review, or approval of the manuscript (all authors)

## Notes

### Competing Interest Statement

The authors have declared no competing interest.

### Funding Statement

This study was funded by the U.S. Food and Drug Administration's Sentinel Initiative. All investigators were supported on FDA WO2006. Dr. Walsh is also supported in part by NIMH
R01MH121455 and R01MH116269.

### Author Declarations

The IRB of Vanderbilt University Medical Center gave ethical approval for this work.

## References

1. Ball, R., Robb, M., Anderson, S. & Dal Pan, G. The FDA’s sentinel initiative—A comprehensive approach to medical product surveillance. Clin. Pharmacol. Ther. 99, 265– 268 (2016).

2. Behrman, R. E. et al. Developing the Sentinel System — A National Resource for Evidence Development. N. Engl. J. Med. 364, 498–499 (2011).

3. Robb, M. A. et al. The US Food and Drug Administration’s Sentinel Initiative: Expanding the horizons of medical product safety. Pharmacoepidemiol. Drug Saf. 21, 9–11 (2012).

4. Platt, R. et al. The FDA Sentinel Initiative — An Evolving National Resource. N. Engl. J. Med. 379, 2091–2093 (2018).

5. Feng, C., Le, D. & McCoy, A. B. Using Electronic Health Records to Identify Adverse Drug Events in Ambulatory Care: A Systematic Review. Appl. Clin. Inform. 10, 123–128 (2019).

6. Liu, F., Jagannatha, A. & Yu, H. Towards Drug Safety Surveillance and Pharmacovigilance: Current Progress in Detecting Medication and Adverse Drug Events from Electronic Health Records. Drug Saf. 42, 95–97 (2019).

7. Fernandes, M. et al. Clinical Decision Support Systems for Triage in the Emergency Department using Intelligent Systems: a Review. Artif. Intell. Med. 102, 101762 (2020).

8. Panahiazar, M., Taslimitehrani, V., Pereira, N. L. & Pathak, J. Using EHRs for Heart Failure Therapy Recommendation Using Multidimensional Patient Similarity Analytics. Stud. Health Technol. Inform. 210, 369–373 (2015).

9. Zhang, P., Wang, F., Hu, J. & Sorrentino, R. Towards personalized medicine: leveraging patient similarity and drug similarity analytics. AMIA Jt. Summits Transl. Sci. Proc. AMIA Jt. Summits Transl. Sci. 2014, 132–136 (2014).

10. Health, C. for D. and R. Postmarket Surveillance Under Section 522 of the Federal Food, Drug, and Cosmetic Act. U.S. Food and Drug Administration https://www.fda.gov/regulatory-information/search-fda-guidance-documents/postmarket-surveillance-under-section-522-federal-food-drug-and-cosmetic-act (2022).

11. Alomar, M., Tawfiq, A. M., Hassan, N. & Palaian, S. Post marketing surveillance of suspected adverse drug reactions through spontaneous reporting: current status, challenges and the future. Ther. Adv. Drug Saf. 11, 2042098620938595 (2020).

12. Bate, A. & Evans, S. J. W. Quantitative signal detection using spontaneous ADR reporting. Pharmacoepidemiol. Drug Saf. 18, 427–436 (2009).

13. Methods | Sentinel Initiative. https://www.sentinelinitiative.org/methods-data-tools/methods.

14. Banerji, A. et al. Natural Language Processing Combined with ICD-9-CM Codes as a Novel Method to Study the Epidemiology of Allergic Drug Reactions. J. Allergy Clin. Immunol. Pract. 8, 1032–1038.e1 (2020).

15. Bayramli, I. et al. Predictive structured-unstructured interactions in EHR models: A case study of suicide prediction. NPJ Digit. Med. 5, 15 (2022).

16. Borjali, A. et al. Natural language processing with deep learning for medical adverse event detection from free-text medical narratives: A case study of detecting total hip replacement dislocation. Comput. Biol. Med. 129, 104140 (2021).

17. Xie, F. et al. Deep learning for temporal data representation in electronic health records: A systematic review of challenges and methodologies. J. Biomed. Inform. 126, 103980 (2022).

18. Sun, W., Rumshisky, A. & Uzuner, O. Evaluating temporal relations in clinical text: 2012 i2b2 Challenge. J. Am. Med. Inform. Assoc. 20, 806–813 (2013).

19. Viani, N. et al. A natural language processing approach for identifying temporal disease onset information from mental healthcare text. Sci. Rep. 11, 757 (2021).

20. Sheikhalishahi, S. et al. Natural Language Processing of Clinical Notes on Chronic Diseases: Systematic Review. JMIR Med. Inform. 7, e12239 (2019).

21. Zech, J., Husk, G., Moore, T., Kuperman, G. J. & Shapiro, J. S. Identifying homelessness using health information exchange data. J. Am. Med. Inform. Assoc. JAMIA 22, 682–687 (2015).

22. Moore, T. et al. Event detection: a clinical notification service on a health information exchange platform. AMIA Annu. Symp. Proc. AMIA Symp. 2012, 635–642 (2012).

23. Bejan, C. A. et al. Mining 100 million notes to find homelessness and adverse childhood experiences: 2 case studies of rare and severe social determinants of health in electronic health records. J. Am. Med. Inform. Assoc. JAMIA 25, 61–71 (2018).

24. Dorr, D. et al. Identifying Patients with Significant Problems Related to Social Determinants of Health with Natural Language Processing. Stud. Health Technol. Inform. 264, 1456–1457 (2019).

25. Desai, R. J. et al. Broadening the reach of the FDA Sentinel system: A roadmap for integrating electronic health record data in a causal analysis framework. NPJ Digit. Med. 4, 170 (2021).

26. Carrell, D. S. et al. Improving Methods of Identifying Anaphylaxis for Medical Product Safety Surveillance Using Natural Language Processing and Machine Learning. Am. J. Epidemiol. 192, 283–295 (2023).

27. Bejan, C. A. et al. Improving ascertainment of suicidal ideation and suicide attempt with natural language processing. Sci. Rep. 12, 15146 (2022).

28. Danciu, I. et al. Secondary use of clinical data: the Vanderbilt approach. J. Biomed. Inform. 52, 28–35 (2014).

29. Walsh, C. G. et al. Prospective Validation of an Electronic Health Record–Based, Real-Time Suicide Risk Model. JAMA Netw. Open 4, e211428 (2021).

30. Wilimitis, D. et al. Integration of Face-to-Face Screening With Real-time Machine Learning to Predict Risk of Suicide Among Adults. *JAMA Netw*. Open 5, e2212095 (2022).

31. Mikolov, T., Sutskever, I., Chen, K., Corrado, G. S. & Dean, J. Distributed Representations of Words and Phrases and their Compositionality. in Advances in Neural Information Processing Systems vol. 26 (Curran Associates, Inc., 2013).

32. Devlin, J., Chang, M.-W., Lee, K. & Toutanova, K. BERT: Pre-training of Deep Bidirectional Transformers for Language Understanding. Preprint at 10.48550/arXiv.1810.04805 (2019).

33. WHO | International Classification of Diseases. WHO http://www.who.int/classifications/icd/en/ (2017).

34. Swain, R. S. et al. A systematic review of validated suicide outcome classification in observational studies. Int. J. Epidemiol. 48, 1636–1649 (2019).

35. Embi, P. J. Algorithmovigilance—Advancing Methods to Analyze and Monitor Artificial Intelligence–Driven Health Care for Effectiveness and Equity. *JAMA Netw*. Open 4, e214622 (2021).

36. Lenert, M. C., Matheny, M. E. & Walsh, C. G. Prognostic models will be victims of their own success, unless…. J. Am. Med. Inform. Assoc. 26, 1645–1650 (2019).

37. Using Population Descriptors in Genetics and Genomics Research: A New Framework for an Evolving Field. (National Academies Press, 2023). doi:10.17226/26902.

38. Viani, N. et al. Annotating Temporal Relations to Determine the Onset of Psychosis Symptoms. Stud. Health Technol. Inform. 264, 418–422 (2019).

39. Ayre, K. et al. Developing a Natural Language Processing tool to identify perinatal self-harm in electronic healthcare records. PloS One 16, e0253809 (2021).

40. Fu, J. T., Sholle, E., Krichevsky, S., Scandura, J. & Campion, T. R. Extracting and classifying diagnosis dates from clinical notes: A case study. J. Biomed. Inform. 110, 103569 (2020).

41. Jin, Y., Li, F., Vimalananda, V. G. & Yu, H. Automatic Detection of Hypoglycemic Events from the Electronic Health Record Notes of Diabetes Patients: Empirical Study. JMIR Med. Inform. 7, e14340 (2019).

42. Cade, B. E. et al. Sleep apnea phenotyping and relationship to disease in a large clinical biobank. JAMIA Open 5, ooab117 (2022).

43. Chen, W., Kowatch, R., Lin, S., Splaingard, M. & Huang, Y. Interactive Cohort Identification of Sleep Disorder Patients Using Natural Language Processing and i2b2. Appl. Clin. Inform. 6, 345–363 (2015).

